# Abnormal gastric electrophysiology following laparoscopic sleeve gastrectomy and associations with symptoms and quality of life

**DOI:** 10.1101/2025.03.10.25323701

**Authors:** Tim Hsu-Han Wang, Chris Varghese, Sam Robertson, Grant Beban, Nicholas Evennett, Daphne Foong, Vincent Ho, Christopher N. Andrews, Stefan Calder, Armen Gharibans, Gabriel Schamberg, Greg O’Grady

**Affiliations:** Department of Surgery, the University of Auckland, New Zealand; Department of Surgery, Auckland City Hospital, Auckland, New Zealand; Alimetry Ltd, Auckland, New Zealand; School of Medicine, Western Sydney University, Sydney, New South Wales, Australia; Department of Gastroenterology and Hepatology, Campbelltown Hospital, Australia; Division of Gastroenterology and Hepatology, University of Calgary, Calgary, Alberta, Canada; Auckland Bioengineering Institute, The University of Auckland, New Zealand

**Author notes:** **Corresponding Author:** Professor Greg O’Grady, Department of Surgery, University of Auckland, New Zealand, Private Bag 92019, Auckland Mail Centre, Auckland 1142, New Zealand.

**Keywords:** Sleeve gastrectomy, gastric myoelectrical activity, high-resolution electrogastrography, slow waves, Gastric Alimetry, body surface gastric mapping

## Abstract

**Background:** Sleeve gastrectomy is an effective bariatric procedure, however may lead to persistent symptoms without obvious mechanical cause. The normal gastric pacemaker region, which lies on the greater curvature of the corpus, is resected in sleeve gastrectomy, however, the electrophysiological consequences are not adequately defined. This study assessed these impacts and associations with symptoms and quality of life (QoL), using non-invasive gastric mapping.

**Methods:** Patients with previous sleeve gastrectomy underwent body surface gastric mapping (Gastric Alimetry, New Zealand), comprising 30-minute fasting baseline and 4-hr post-prandial recordings. Analysis encompassed Principal Gastric Frequency (PGF), BMI-adjusted amplitude, Gastric Alimetry Rhythm Index (GA-RI), with comparison to reference intervals and matched controls. Symptoms were evaluated using a validated App and questionnaires.

**Results:** 38 patients (median 36 months post-surgery; range 6-119 months) and 38 controls were recruited. 35/38 patients had at least one abnormal parameter, typically reduced frequencies (2.3±0.34 vs controls 3.08±0.21; p<0.001) and amplitudes (14.8±6.9 vs 31.5±17.8; p<0.001). Patients exhibited higher symptoms and lower QoL (PAGI-SYM 20 vs controls 7, p<0.001; PAGI-QOL 27 vs 136, p<0.001). Gastric amplitude and GA-RI correlated positively with bloating (r=0.71, p<0.001 and r=0.60, p=0.02) while amplitude correlated negatively with heartburn (r=-0.46, p=0.03). Lower gastric amplitudes also correlated with greater weight loss (r=-0.45; p=0.014).

**Conclusion:** Sleeve gastrectomy modifies gastric electrophysiology due to pacemaker resection, with variable remodelling. Substantial reductions in gastric frequency and amplitude occur routinely after surgery, and specific relationships between post-sleeve gastric amplitude, symptoms of heartburn and bloating, and weight loss are identified.

## Introduction

Sleeve gastrectomy is a common procedure for obesity and its associated consequences including diabetes and metabolic syndrome. A significant portion of the greater curvature is resected from antrum to fundus, reducing gastric volume (1). While most people experience excellent outcomes in terms of weight loss, metabolic outcomes and quality of life (QoL) (2), up to 30% develop persistent foregut symptoms such as nausea, post-prandial bloating, heartburn and reflux (3–7). These symptoms are commonly evaluated endoscopically and radiologically and are usually managed medically. However, a subset of patients suffer chronic symptoms despite normal investigations and no evident mechanical cause. These symptoms may necessitate conversion (e.g. to gastric bypass), and increase morbidity and costs of care (8, 9).

Gastric motility is controlled by a gastric pacemaker region located at the greater curvature of the upper corpus, from which slow waves propagate circumferentially then antegrade, at a frequency close to 3 cycles per minute (cpm), before rapidly accelerating in the terminal antrum (10–13). This normal pacemaker region is completely resected during a sleeve gastrectomy procedure, however, the implications for gastric motility and post-surgical symptoms remain poorly understood (14, 15).

A new non-invasive technique for monitoring gastric motility has recently emerged, called body surface gastric mapping, with a commercial device called Gastric Alimetry (Alimetry, New Zealand) recently achieving clinical translation and regulatory clearances (16, 17). This new test also includes an App allowing simultaneous validated symptom profiling, enabling robust symptom correlation with gastric electrical abnormalities (17). Recent studies using this technology are promising, revealing a range of slow wave initiation and conduction abnormalities in medical and surgical conditions (18–24).

This study therefore aimed to assess the long-term effects of sleeve gastrectomy on gastric function measured by BSGM, and their contributions to persistent gastric symptoms and postoperative quality of life (QoL).

## Methods

Ethical approval for this study was granted by the institutional review committees of the University of Auckland, Western Sydney University and The University of Calgary Conjoint Health Research Ethics Board (AH1125, H15157, REB19-1925). Consecutive eligible patients who underwent a sleeve gastrectomy within the past 10 years were then recruited from these institutions. Patients were excluded if they had a history of skin allergy to adhesives, evidence of mechanical gastric or small bowel obstruction as a cause for their symptoms. Informed consent was obtained from all patients. A healthy control cohort was also recruited having no significant history of gastric disorders, gastric symptoms, gastrointestinal procedures, diabetes, or medications affecting gastrointestinal function.

### Body Surface Gastric Mapping

Gastric Alimetry was performed under a protocol adapted for patients who had a reduced gastric volume. This device comprises a high-resolution (HR) stretchable electrode array (8×8 electrodes; 20 mm spacing; 196 cm^2^), a wearable Reader, validated iOS app for symptom logging, and a cloud-based reporting platform (**Figure 1A,B**) (25–27). Baseline recordings were performed in the first 30 minutes, followed by a 218 kCal meal, comprising 100mL of Ensure (93kCal; Abbott Nutrition, IL, USA) and half an oatmeal energy bar (125kcal, 2.5g fat, 22.5g carbohydrate, 5g protein, 3.5g fibre; Clif Bar & Company, CA, USA), consumed over 10 minutes, and a 4-hr postprandial recording in order to capture a full gastric activity cycle. Patients were seated in a reclined chair and were asked to limit movement, talking, and sleeping, but were able to read, watch media, work on a mobile device, and mobilize for comfort breaks (28). Controls underwent the standard Gastric Alimetry test using the standard 482kCal meal (28). The impact of different test meal sizes between the two groups is known, having been rigorously analysed in a separate study quantifying modest reductions in post-prandial amplitudes and rhythm index with smaller meal sizes (refer discussion) (29).

**Figure 1:**
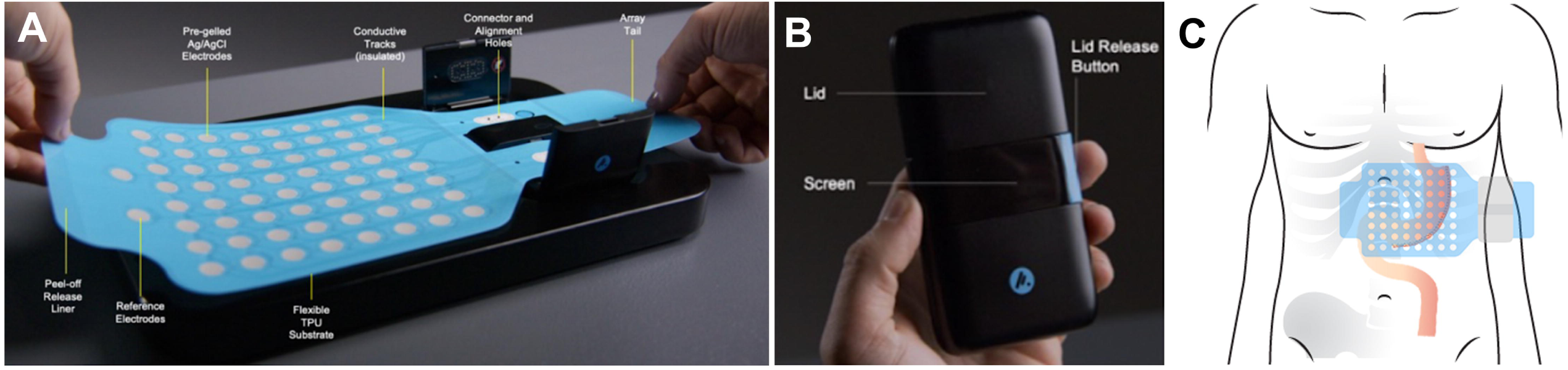
A. Photographic representation of the Gastric Alimetry array. B. Photograph of the Gastric Alimetry reader. C. Representative diagram of the sleeve gastrectomy and its relation to array placement.

Spectral analysis was performed, encompassing the four established test metrics (30): Principal Gastric Frequency (PGF; cycles per minute, cpm), BMI-Adjusted Amplitude (µV), Gastric Alimetry Rhythm Index (GA-RI; reflecting pacemaker stability), and fed:fasted Amplitude Ratio (ff-AR; indicating meal response with contractions). The metrics were assessed using published normal ranges (31). Frequency was not reported if stable gastric rhythm was not detected (26). Adverse events were recorded.

### Symptom and Quality of Life Assessments

Symptom development during the test was recorded on the validated Gastric Alimetry App and summarized as a Total Symptom Burden Score (TSBS) (27). Symptoms and QoL over the preceding two weeks were further assessed using validated questionnaires, comprising the Patient Assessment of Gastrointestinal Symptoms (PAGI-SYM), Patient Assessment of Gastrointestinal Quality of Life (PAGI-QOL) and European Quality of Life 5 Dimension (EQ-5D-5L) (32–34).

### Data Analyses

Statistical analysis was performed using GraphPad Prism (San Diego, CA, USA) and R v.4.0.1 (R Foundation for Statistical Computing, Vienna, Austria). Spectral analysis comparisons with the age and gender matched healthy control patients were performed using the unpaired Student’s t-test. All data is presented as mean +/- standard deviation. Correlation analyses were then performed between the test spectral metrics, symptoms, and QoL scores, using Spearman’s correlation. A threshold of p<0.05 was deemed to be statistically significant.

## Results

38 patients were recruited including 26 females of median age 51.7 years (range 22-73 years), with a median BMI 33.0 (range 20.4-47.3). The median duration since surgery was 46.2 months (range 6-119 months). Indications for all sleeve gastrectomy were obesity. Patient and matched control demographics are presented in **Table 1**, with controls being of moderately lower BMI (33.0 ±6.0 vs 27.4±8.0; p<0.001) and slightly lower age (51.7±11.8 vs 44.0±14.6; p=0.01), noting that Gastric Alimetry metrics are BMI-adjusted (30).

**Table 1:** Patient and match control demographic, quality of life (QOL) and spectral characteristics data. The p values in bold denote statistically significant difference between the groups. TSBS- Total Symptom Burden Score. PAGI-SYM- Patient Assessment of Upper Gastrointestinal Symptom Severity Index. PAGI-QOL- Patient Assessment of Upper GastroIntestinal Disorders-Quality of Life. EQ-5D-5L- questionnaire developed by the EuroQol Group. ff-AR- fed:fasted Amplitude Ratio. GA-RI- Gastric Alimetry Rhythm Index.

| **Characteristics** | **Patients**<br>**(n=38)** | **Matched controls**<br>**(n=38)** | **p**<br>**value** |
| --- | --- | --- | --- |
| Age | 51.7 ± 11.8 | 44.0 ± 14.6 | 0.01 |
| BMI (kg/m<sup>2</sup>) | 33.0 ± 6.0 | 27.4 ± 8.0 | <0.001 |
| Gender |  |  |  |
| Male | 12 | 11 | 1 |
| Female | 26 | 27 |  |
| Duration since surgery<br>(months) | 46.2 ± 32.8 | N/A |  |
| **QOL** |  |  |  |
| TSBS | 5.9 ± 7.4 | 2.7 ± 7.0 | 0.06 |
| PAGI-SYM | 20 ± 19 | 7 ± 14 | <0.001 |
| PAGI-QOL | 27 ± 29 | 136 ± 20 | <0.001 |
| EQ-5D-5L | 0.86 ± 0.19 | 0.96 ± 0.09 | <0.01 |
| Self-reported EQ-5D-5L | 74 ± 21 | 88 ± 8 | <0.001 |
| **Spectral Characteristics** |  |  |  |
| Frequency (2.65 - 3.35cpm) | 2.40 ± 0.22 | 3.09 ± 0.21 | <0.001 |
| Adjusted amplitude (22-70μV) | 28.2 ± 7.1 | 38.8 ± 15.3 | <0.001 |
| ff-AR (≥1.08) | 1.54 ± 0.51 | 1.99 ± 0.91 | 0.01 |
| GA-RI (≥0.25) | 0.33 ± 0.14 | 0.50 ± 0.21 | <0.001 |

### HR Gastric Mapping

Of the 38 patients, 35 had at least one abnormal gastric slow wave activity parameter compared to the normative test reference range (**Table 1**). Of the 24 patients with a detectable Principal Gastric Frequency, 21 were found to be reduced (mean 2.30 cpm), while the remaining 14 patients had an undetectable frequency due to either very irregular gastric rhythms, or inability to detect remnant gastric activity cutaneously. 12/38 patients had 2 abnormal gastric activity characteristics, including a combination of abnormal PGF with low GA-RI (n=3), ff-AR (n=8) and BMI-adjusted amplitude (n=4). Overall, 4/38 patients were found to have reduced BMI-adjusted amplitude, 8/38 had reduced ff-AR, 10/38 had reduced GA-RI. An example spectral map of a healthy control participant is shown in **Figure 2**, together with normative reference interval ranges, while two typical examples of patient spectral maps are shown in **Figure 3**.

**Figure 2:**
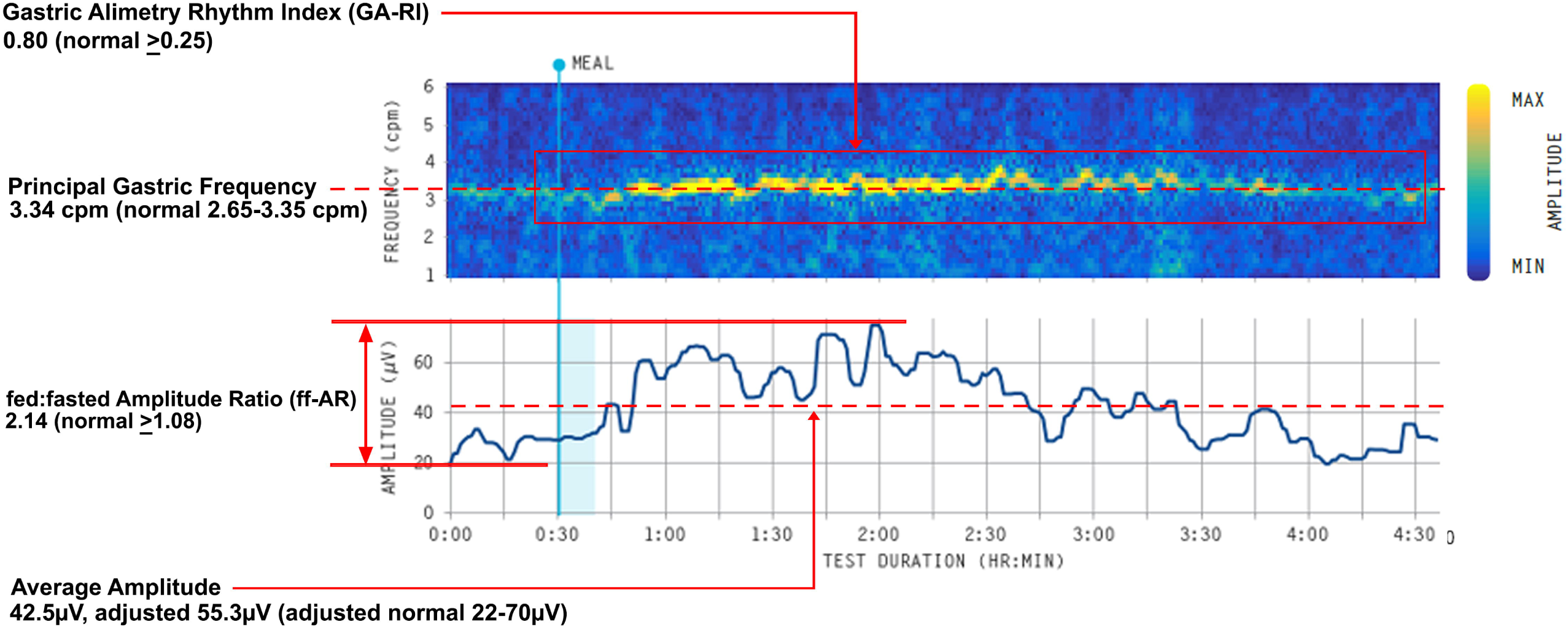
Example spectral diagram of a healthy matched control case. Normative values are provided in brackets.

**Figure 3:**
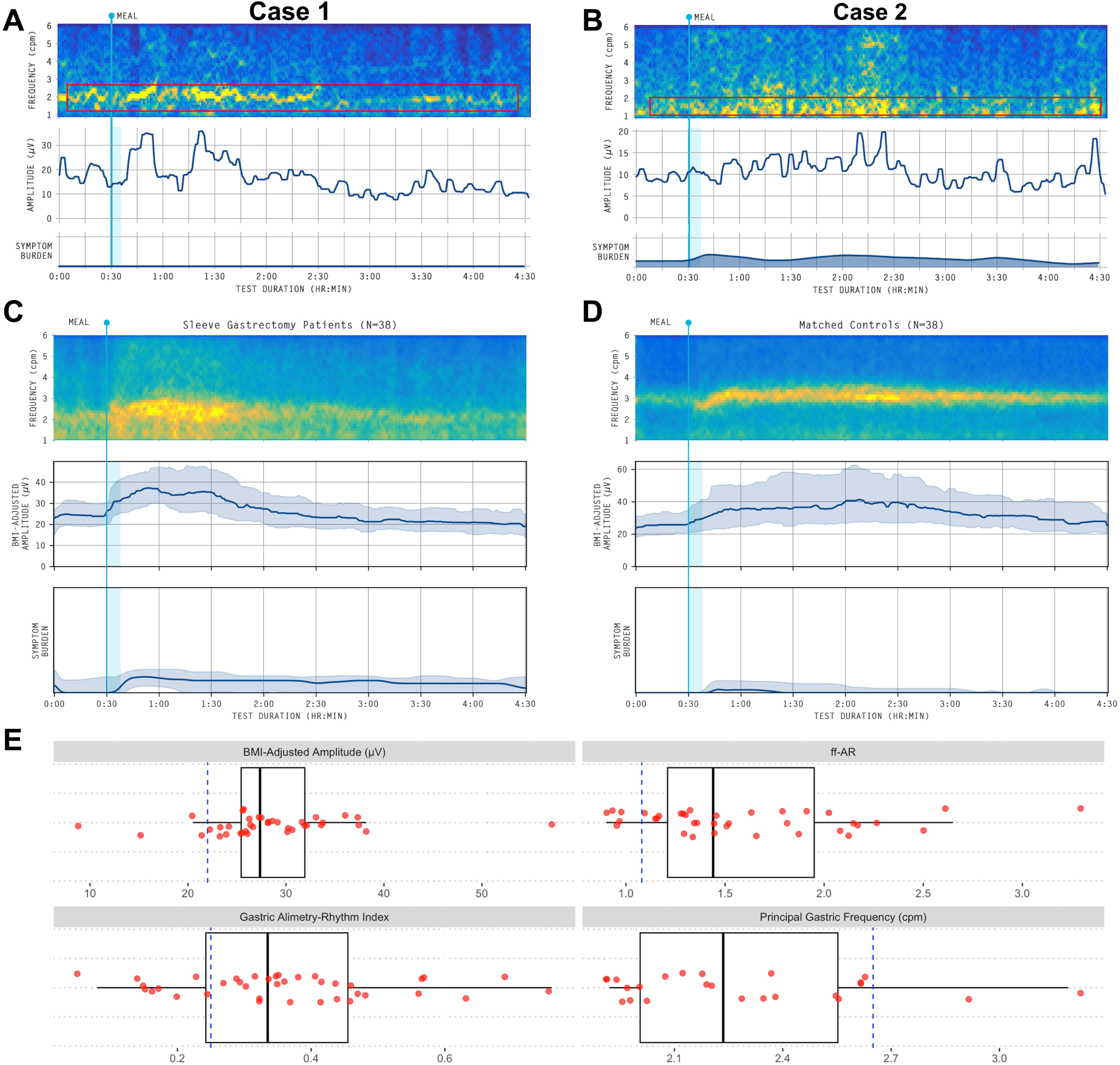
Example spectral diagram of cases from A. Patient with stable low frequency gastric activity B. Patient with unstable gastric activity with low amplitude and moderate symptom development. Boxed red line in A, B indicate the degraded principal gastric frequency, however, these are significantly lower in amplitude than healthy patients. C, D are average spectrogram with median (IQR shaded) BMI-adjusted amplitude and symptom burden for sleeve gastrectomy cohort (C), and matched control cohort (D). E. Box and whisker diagram of Principal Gastric Frequency, BMI-adjusted amplitude, GA-RI and ff-AR. Dashed blue line denotes the lower limit of normal range.

When the pooled results were compared to the matched cohort, the patient cohort was found to be statistically different in all gastric slow wave characteristics including frequency (2.30 vs 3.08 cpm; p<0.001), BMI-adjusted amplitude (28.4 vs 31.5 µV; p<0.001), ff-AR (1.60 vs 2.00; p=0.02) and GA-RI (0.35 vs 0.50; p<0.001) (**Table 1**, and averaged spectrogram data for visual whole-cohort comparisons presented in **Figure 3C,D**). Visual analysis of the averaged spectrograms of patients vs controls also demonstrates how gastric pacemaker rhythm became significantly more variable after sleeve gastrectomy, with a wider, shorter and more irregular gastric spectral activity band (**Figure 3C vs 3D**).

No adverse events occurred due to BSGM testing.

### QoL, Symptom and Weight Loss Evaluations

The QoL data and TSBS are presented in **Table 1**. Overall, there were 14/38 patients who were found to have significant symptoms during the test, judged by having a TSBS ≥5, of which 7/14 had substantial symptoms with TSBS≥10. Overall, the TSBS result in the patient cohort was 5.9 compared to 2.6 in matched controls (p=0.06). The patient cohort also showed higher symptom burdens and substantially reduced QOL compared to matched controls in the other instruments, including PAGI-SYM (20 vs 7; p<0.001), PAGI-QoL (27 vs 136; p<0.001), EQ-5D-5L (0.86 vs 0.96; p<0.01) and self-reported EQ-5D-5L (74 vs 88; p<0.001).

**Table 2** reports correlation analyses between the gastric mapping parameters and individual symptom measures. BMI-adjusted amplitude was positively correlated with bloating (r=0.71, p<0.001) but inversely correlated with heartburn (r= 0.46, p=0.03). GA-RI likewise showed a significant positive correlation with bloating (r=0.60, p=0.002). No other significant correlations were observed between the remaining mapping metrics and symptom indices. Taken together, these results indicate that heightened gastric slow-wave activity post-sleeve gastrectomy is associated with bloating, while reduced amplitude may predispose to heartburn.

**Table 2:** Pearson correlations between patient-reported symptoms and Gastric Alimetry metrics

| Symptom | Metric | R value | p value |
| --- | --- | --- | --- |
| Bloating | BMI-Adjusted Amplitude | 0.71 | <0.001 |
| Heartburn | BMI-Adjusted Amplitude | -0.46 | 0.03 |
| Bloating | GA-RI | 0.60 | 0.002 |

Additionally, lower BMI-adjusted amplitudes at the time of mapping were associated with greater weight reduction from time of referral (r=-0.45; p=0.014), whereas frequency was not correlated with change in weight (p=0.25) (**Figure 4**).

**Figure 4:**
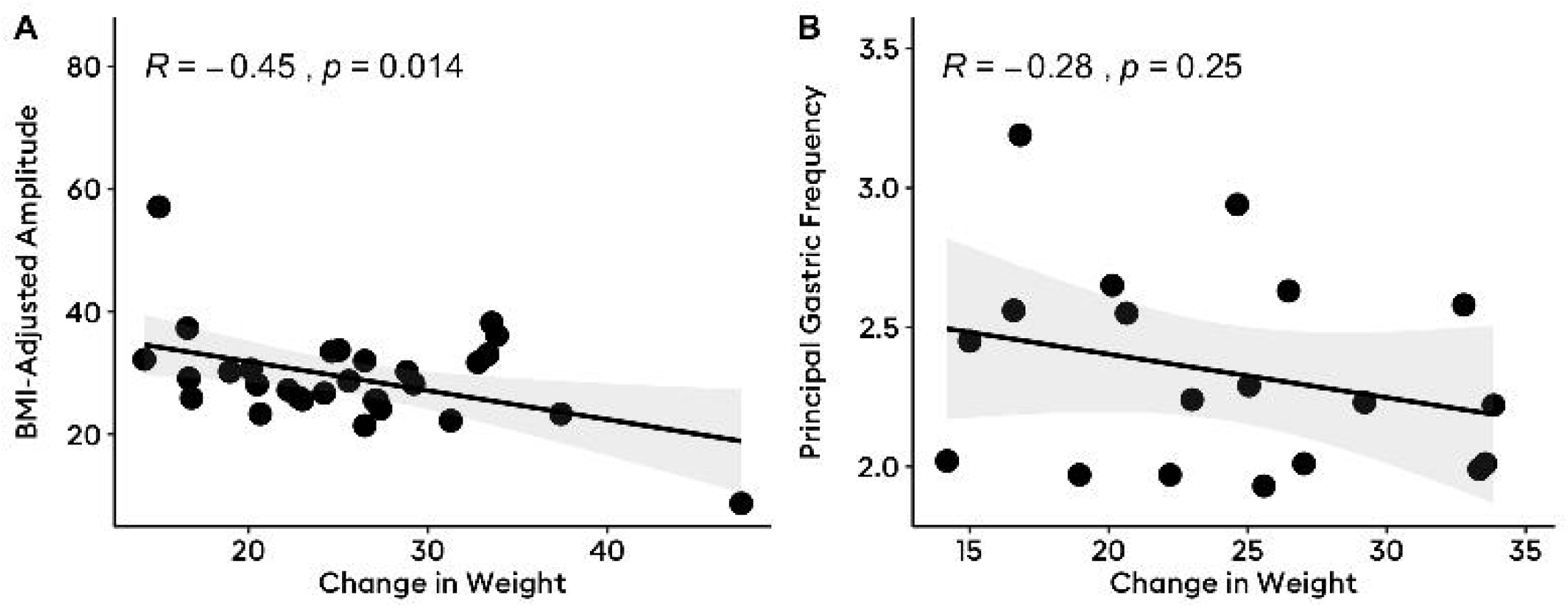
Pearson correlation between body surface gastric mapping BMI-Adjusted Amplitude (A) and Principal Gastric Frequency (B) and change in weight from referral to follow-up.

## Discussion

In this body surface gastric mapping study investigating the electrical activity of the stomach following a sleeve gastrectomy, our results showed consistent changes in gastric electrophysiology occur after sleeve gastrectomy. Almost all patients were found to have substantially reduced gastric frequencies, and a further 59% having one or more additional gastric electrical abnormalities in amplitude or rhythm. All metrics were substantially lower than observed in the matched controls. Specific relationships between post-sleeve electrophysiology and symptoms of heartburn and bloating, as well as weight loss, were also identified.

Sleeve gastrectomy is one of the most widely performed bariatric procedures. While most patients experience excellent outcomes, some develop persistent post-operative symptoms including reflux, nausea, pain and excessive food intolerance. While some chronic symptoms are attributable to mechanical causes (such as stenosis or twisting of the gastric sleeve), a significant portion of these patients have unexplained symptoms. Previous studies have shown that gastric electrical abnormalities can arise after sleeve gastrectomy (14, 15, 35) secondary to resection of the native gastric pacemaker. This study extends these findings in demonstrating consistent degradation of the intrinsic pacemaker frequency, reduced amplitude, and meal-responses robustly associated with gastric symptoms. This investigation was enabled through non-invasive body surface gastric mapping, extending insights from smaller cohorts of patients that had undergone invasive intraoperative mapping (14).

Early investigations into the surgical effects of disconnecting the normal gastric pacemaker region were conducted using canine models in the early 1970s (36, 37). In one classic study, Kelly and Code performed a longitudinal bisection of the canine stomach and observed the gastric electrical activity on either side of the bisection using sparse implanted electrodes. Abnormal gastric electrical activity was observed on the lesser-curvature half post-operatively, at a reduced frequency and with irregular pacemaking. Intrinsic frequency gradients have also been identified longitudinally in the stomach from corpus to antrum, and transversely from greater to lesser curvature, such that loss of the normal dominant highest-frequency pacemaker leads to breakout of subordinate, lower frequency, and less-stable pacemaker sites nearer to the lesser curvature (12, 38). These gradients are related to denser ICC networks occurring along the greater curve, which are resected during sleeve gastrectomy (12). Our results are consistent with these established physiological data, mostly from classic animal studies, as it similarly revealed emergence of lower-frequency pacemaking of reduced stability in the human gastric sleeve remnant.

The only previous high-resolution mapping study of sleeve gastrectomy was performed by Berry *et al*, where 8 patients had invasive intra-operative mapping of the stomach before and after sleeve gastrectomy (14). In that study, abnormal gastric electrical activity was observed, including the presence of retrograde gastric slow wave activity. Berry et al performed gastric mapping immediately after the resection, and were therefore limited by the invasive nature of the method, meaning small numbers of patients were studied, and gastric recovery and symptom correlations could not be evaluated. Another notable recent case series was performed by Gharibani *et al*, where 20 patients who had a laparoscopic sleeve gastrectomy were studied (35). Here, low-resolution electrogastrography (EGG) was performed, with the authors also finding a reduced frequency of gastric pacemaking activity. Through these and other studies (15), it is consistently seen that there are significant changes to the gastric conduction system following sleeve gastrectomy. These conduction abnormalities are now robustly expanded through state-of-the-art gastric mapping techniques in the current work, together with symptom correlations.

The major advantage of the non-invasive Gastric Alimetry system is that gastric activity can be recorded reliably cutaneously, with major improvements compared to legacy EGG techniques (39). The system also allows for the simultaneous measurement of symptoms during the test (27). In the current study, the evaluations were limited to spectral parameters (frequency, BMI-adjusted amplitude and rhythm index), however, in future it may also be possible to assess slow wave propagation direction non-invasively using body surface gastric mapping (40, 41). This would allow to measure the directionality of slow wave propagation, which is of interest given previous reports of retrograde electrical propagation invasive serosal mapping studies after surgical alterations, including breakout events in the remnant antrum after sleeve gastrectomy (14), and retrograde propagation from the higher intrinsic frequency of the duodenum after a gastroduodenal anastomosis post antrectomy (21). Gharibans *et al* have previously shown that retrograde propagation can be associated with dyspeptic symptoms and nausea (40), and disorganised propagation could also be associated with reflux, which is a major complication of sleeve gastrectomy (42).

Notably, the gastric slow wave amplitude in our patient cohort was also substantially lower than the controls, despite adjusting for BMI. This is a novel finding and likely reflects resection of a large mass of ICC of smooth muscle tissue, such that reduced ionic current flows ultimately arrive at the skin surface. However, a limitation should be noted in that sleeve gastrectomy patients were also only able to eat a smaller size meal, which likely also partly account for some, but not all, of the group-level amplitude reduction (29). Of note, lower BMI-adjusted amplitude was also associated with greater weight reduction, possibly reflecting the fact that greater gastric resections resulted in a reduced functional sleeve capacity.

This study found that patients who had undergone a sleeve gastrectomy had significantly worse QoL compared to the matched cohort in all questionnaires employed. Sleeve gastrectomy generally results in an improved QoL vs patients’ baselines (2, 43, 44), although there are also studies indicating that a significant proportion of post-operative patients have QoL compromised by persistent symptoms (45). More recent studies have now found a return to baseline QoL results in patients who are followed-up longer-term (46). Our cohort appeared to have a relatively high symptom burden, which could also be in-part due to channelling bias in our cohort, where patients with persistent symptoms would be more likely to participate in research.

Our findings indicate that altered gastric slow-wave amplitude, rather than frequency, may contribute to persistent symptoms some patients experience after sleeve gastrectomy. Specifically, higher BMI-adjusted amplitude, associated with a more stable pacemaker rhythm (GA-RI), were associated with bloating; whereas reduced amplitude correlated with increased heartburn. These relationships offer an electrophysiological explanation for postoperative symptom burden that extends beyond purely mechanical or anatomical causes, and point to the emerging role for Gastric Alimetry in clarifying symptom genesis after gastric surgery (24). Moreover, it is plausible that patients with certain predispositions—such as diabetic gut dysfunction (46)—are at greater risk of developing disordered post-sleeve motility, and may therefore benefit from preoperative functional screening to guide the choice of bariatric procedure. Gastric Alimetry could therefore be incorporated as a tool for patient selection, for instance favouring gastric bypass in cases where preoperative mapping demonstrates marked dysmotility. Looking ahead, future research may also explore how targeted interventions—such as external gastric pacing or ablation of anomalous pacemaker sites (47, 48)—might address pathologic gastric rhythms within the sleeve remnant and ultimately improve long-term symptom control.

Despite extensive attempts to recruit a well-matched control cohort, ultimately some minor differences were apparent in age and BMI. To mitigate this, all data were also compared to a normative reference range devised from 110 healthy controls encompassing a range of BMI, age, sex and ethnicity (31), and Gastric Alimetry metrics were BMI-adjusted (30).

In conclusion, a novel non-invasive gastric mapping device, Gastric Alimetry, was used to assess gastric electrical activity and symptoms following sleeve gastrectomy. Our results show that remodelling and recovery of the gastric pacemaker system is variable, with reduced gastric pacemaker frequency, reduced pacemaker stability, and weaker contractile amplitudes being characteristic after sleeve gastrectomy. Symptom correlations with gastric activity clarify some of the symptom associations that may follow sleeve gastrectomy, and point to the emerging role of non-invasive electrophysiology tools in post-surgical diagnostics.

## Data Availability

All data produced in the present study are available upon reasonable request to the authors

## Acknowledgements

We thank the volunteers who participated in this research and our Auckland clinical research coordinators Gen Johnston and India Wallace. We also thank Elaine Yi, Auckland City Hospital Upper Gastrointestinal Surgery clinical nurse specialist, for assisting with patient recruitment.

## Funding

This work and authors were supported by the New Zealand Health Research Council, The Royal Australasian College of Surgeons John Mitchell Crouch Fellowship (GOG), the National Institutes of Health (R56 126935), and the Auckland Medical Research Foundation Douglas Goodfellow Medical Research Fellowship (THHW).

## Notes

### Competing Interest Statement

The authors have declared no competing interest.

### Author Declarations

Ethical approval for this study was granted by the institutional review committees of the University of Auckland, Western Sydney University and The University of Calgary Conjoint Health Research Ethics Board (AH1125, H15157, REB19-1925).

